# Machine Learning Approach to Identify Gut Microbiota Biomarkers in Patients With ST-Elevation Myocardial Infarction Presenting With Primary Ventricular Tachyarrhythmias

**DOI:** 10.64898/2026.09.16.26363271

**Authors:** Hsu-Po Tseng, Zi-Lun Lai, Yi-Yao Hsu, Yuan-Hua Hung, Der-Yang Cho, Po-Ren Hsueh, Wei-Hsin Chung, Mei-Yao Wu, Yen-Nien Lin, Ke-Wei Chen, Shih-Sheng Chang, Kuan-Cheng Chang

## Abstract

**Background:** Risk stratification for primary ventricular tachycardia/ventricular fibrillation (VT/VF) in patients with ST-elevation myocardial infarction (STEMI) remains limited. We aimed to identify gut microbiome biomarkers and microbial metabolic pathways associated with primary VT/VF in STEMI using machine learning (ML).

**Methods:** Stool samples from 33 STEMI patients (7 with primary VT/VF and 26 without VT/VF) underwent full-length 16S rRNA sequencing during the acute (≤7 days) and recovery (30–60 days) phases. An ML pipeline integrating taxonomic features identified reproducible features associated with primary VT/VF. Core biomarkers were defined as taxa demonstrating both univariate significance (false discovery rate [FDR] <0.05) and multivariate classification importance. Functional pathways and enzymes were predicted using PICRUSt2.

**Results:** Five acute-phase taxa were reproducibly associated with primary VT/VF: Clostridium aldenense, Enterocloster bolteae, bacterium NLAE-zl-G101, Alistipes shahii, and Roseburia sp. (all FDR <0.05). A reduced-feature support vector machine model achieved an area under the receiver operating characteristic curve of 0.846 (95% confidence interval, 0.676–0.984), with 85.7% sensitivity and 76.9% specificity. Differential abundance analysis showed enrichment of Bacteroides fragilis, Bacteroides thetaiotaomicron, and E. bolteae. Functional analysis demonstrated enrichment of fucose degradation and purine catabolism, with increased predicted abundances of L-fucose mutarotase and xanthine dehydrogenase.

**Conclusions:** Primary VT/VF in STEMI was associated with a distinct gut microbiome–metabolic signature characterized by Lachnoclostridium-related taxa, persistent Bacteroides enrichment, and enhanced predicted fucose and purine metabolism. These exploratory findings warrant validation in larger independent cohorts and mechanistic studies.

**Clinical Perspective:** **What Is New?**

- In this exploratory cohort of 33 patients with STEMI, including 7 with primary VT/VF, an integrated microbiome and machine-learning framework identified five acute-phase microbial taxa associated with primary VT/VF and a coordinated predicted metabolic signature involving fucose and purine pathways.

**What Are the Clinical Implications?**

- Because stool sampling occurred after the arrhythmic event and the VT/VF subgroup was small, these findings should be considered hypothesis-generating rather than predictive; independent prospective validation is required before microbiome-derived markers can be considered for clinical risk stratification.

## Introduction

Sudden cardiac death (SCD) remains a leading cause of mortality in STEMI patients, primarily due to sustained ventricular tachyarrhythmias, such as ventricular tachycardia and ventricular fibrillation (VT/VF).^1^ Within the first 48 hours after symptom onset, 4-12% of patients with STEMI are at risk of primary VT/VF, a statistic significantly exceeding that of NSTEMI patients.^1,2^ These early VTAs in STEMI are critical not only for their association with SCD but also because they are linked to increased mortality both during in-hospital stage and over the long term.^3^ Furthermore, the contributors to the ventricular tachyarrhythmic SCD following STEMI are multi-factorials involving the triggers and the substrate generated secondary to coronary artery occlusion/reperfusion, myocardial ischemia, augmented sympathetic tone, metabolic disturbance, and altered sarcolemma ionic currents.^4^ This complex etiology highlights the need for a comprehensive approach to predict and manage the malignant ventricular tachyarrhythmias.

The human gastrointestinal tract harbors a complex microbial ecosystem consisting of trillions of microorganisms that collectively regulate metabolic, immune, and inflammatory homeostasis. Growing evidence supports a close relationship between the gut microbiome and cardiovascular disease, including STEMI.^5–7^. Several mechanisms have been proposed to mediate this interaction, including increased intestinal permeability, microbial metabolite production, and alterations in microbial community composition.^5,8^ For example, an increased Firmicutes-to-Bacteroidetes ratio has been reported in both STEMI and hypertension.^6^ Although it remains uncertain whether these microbial alterations are merely consequences of cardiac injury or active contributors to disease progression, recent studies support the existence of a bidirectional heart–gut axis. It has been shown that myocardial ischemia–reperfusion injury impairs intestinal barrier integrity, promoting bacterial translocation and exacerbating cardiac inflammation.^5^ Despite these advances, whether gut microbial dysbiosis contributes to the development of life-threatening ventricular arrhythmias during STEMI remains largely unknown.

Accumulating evidence further suggests that specific microbial taxa and their metabolites may influence cardiovascular risk. Compared with healthy individuals, STEMI patients exhibit reduced abundances of beneficial commensals such as *Bifidobacterium* and increased abundances of taxa including *Desulfovibrio* and *Lactobacillus*.^6^ These microbial alterations may affect host physiology through metabolite production. For instance, several short-chain fatty acid (SCFA)-producing taxa, including *Prevotella*, members of the *Lachnospiraceae* family, Eubacterium *rectale*, *Lactobacillales*, and *Bifidobacterium*, have been reported to be depleted in STEMI patients. Because SCFAs exert anti-inflammatory and cardioprotective effects, depletion of these taxa may contribute to the pathophysiology of STEMI.^9^ Together, these findings suggest that alterations in microbial composition and metabolic function may influence cardiovascular pathophysiology beyond traditional risk factors.

While the gut microbiome has emerged as a potential contributor to STEMI pathogenesis through immune modulation, metabolic signaling, and endothelial dysfunction, its role in the development of primary VT/VF in STEMI remains poorly understood. Moreover, no microbiome-based strategy has been established for risk stratification of ventricular arrhythmic complications in STEMI. Therefore, this study aimed to characterize differences in gut microbial composition between STEMI patients with and without primary VT/VF using a machine learning–based analytical framework and investigate associated functional metabolic pathways through computational microbiome profiling.

## Methods

### Study Cohort and Sample Collection

This study was conducted in accordance with the Declaration of Helsinki and was approved by the Institutional Review Board of China Medical University Hospital (CMUH111-REC3-195). Written informed consent was obtained from all participants prior to enrollment. Fecal samples and corresponding clinical data were collected from patients admitted to CMUH between February 1, 2023, and December 31, 2024. STEMI was diagnosed according to contemporary guideline criteria based on characteristic electrocardiographic findings and confirmation by coronary angiography. Primary VT/VF was defined as ventricular tachycardia or ventricular fibrillation occurring within 48 hours of STEMI onset and requiring emergent electrical defibrillation for hemodynamic stabilization.

### Data Availability Statement

The data that support the findings of this study are available from the corresponding author upon reasonable request, subject to institutional and ethical restrictions governing participant data.

Stool samples were collected at two predefined time points: the acute phase of STEMI (0– 7 days after primary percutaneous coronary intervention [PCI]) and the recovery phase (30–60 days after PCI). The primary analysis compared acute-phase STEMI patients with and without primary VT/VF. Control subjects without known coronary artery disease or a history of STEMI were included for baseline comparisons. Stool samples obtained during the recovery phase were used as longitudinal references to evaluate temporal changes in the gut microbiome following the acute STEMI event.

### Fecal DNA Extraction

Genomic DNA was extracted from homogenized stool samples using the QIAamp PowerFecal Pro DNA Kit (QIAGEN, Hilden, Germany) according to the manufacturer’s instructions. To minimize cross-contamination and ensure procedural consistency, DNA extraction was performed on the automated QIAcube HT platform (QIAGEN). DNA concentration was measured using a Qubit 3.0 Fluorometer with the Qubit dsDNA High Sensitivity Assay Kit (Invitrogen, Carlsbad, CA, USA). DNA quality was assessed prior to library preparation, and all samples were stored at −80°C until further processing.

### Full-Length 16S rRNA Amplification and Sequencing

Full-length bacterial 16S rRNA genes were amplified for PacBio long-read sequencing using universal primers 27F (5′-AgRgTTYgATYMTggCTCAg-3′) and 1492R (5′-RgYTACCTTgTTACgACTT-3′), targeting the V1–V9 hypervariable regions. PCR reactions were performed in 25 μL volumes containing KAPA HiFi HotStart 2× ReadyMix (Kapa Biosystems, Woburn, MA, USA), 0.375 mM of each primer, and 500–1000 ng of template DNA. Thermal cycling consisted of an initial denaturation at 95°C for 3 minutes, followed by 25 cycles of 95°C for 30 seconds, 57°C for 30 seconds, and 72°C for 1 minute, with a final extension at 72°C for 1 minute. Amplicons were verified by 0.8% agarose gel electrophoresis, quantified using the Qubit dsDNA HS Assay, and purified with AMPure PB magnetic beads (PacBio, Menlo Park, CA, USA). SMRTbell libraries were prepared from 500 ng of purified amplicons using the SMRTbell Express Template Prep Kit 2.0 (PacBio). Sequencing was performed on the PacBio Sequel IIe platform using SMRT Cell 8M v3 consumables. Circular consensus sequencing (CCS) reads were generated using SMRT Link v9.0 according to standard PacBio workflows.

### Bioinformatics Analysis

Raw sequencing data were processed using the PacBio HiFi full-length 16S workflow integrating QIIME 2 and DADA2 for amplicon sequence variant (ASV) generation.^14, 15^ Sequence quality control was performed using SeqKit,^10^ followed by primer trimming with Cutadapt. Reads were subsequently denoised,^11^ Reads were subsequently denoised, dereplicated, and screened for chimeras using DADA2 within QIIME 2. Taxonomic assignment was performed using VSEARCH against the SILVA reference database (version 138), generating ASV-level taxonomic feature tables for downstream analyses.^12,13^

### Microbial Diversity Analysis

Alpha diversity was assessed using Observed richness, Chao1, Shannon, and Simpson indices. Differences between groups were evaluated using one-way analysis of variance (ANOVA). Beta diversity was evaluated using unweighted UniFrac, weighted UniFrac, Jaccard, and Bray–Curtis distance metrics. Group differences in microbial community composition were assessed using permutational multivariate analysis of variance (PERMANOVA).

### Taxonomic Biomarker Discovery and Functional Prediction

Differentially abundant taxa were identified using linear discriminant analysis effect size (LEfSe), with a linear discriminant analysis (LDA) score threshold ≥2.0. ^14^ Functional pathway prediction was performed using PICRUSt2^21^ and between-group comparisons of predicted metabolic pathways and enzyme abundances were conducted using STAMP.^22^.

### Machine Learning–Based Biomarker Discovery

To enhance robustness and minimize model-specific bias, microbial features were analyzed through two complementary pipelines before final model construction.

#### 1) Univariate Differential Abundance Analysis

Differential abundance testing was performed in MicrobiomeAnalyst using the edgeR framework with relative log expression normalization and negative binomial modeling.^15^ Multiple testing correction was performed using the Benjamini–Hochberg procedure, and taxa with a false discovery rate (FDR) <0.05 were considered statistically significant.^16^

#### 2) Machine Learning–Based Feature Selection

For classification modeling, count data were transformed using centered log-ratio (CLR) normalization with a pseudocount of 1.17 Feature selection was performed using a bootstrap-based framework comprising 300 stratified bootstrap iterations across 30 independent random seeds. Three complementary feature-selection algorithms were employed: least absolute shrinkage and selection operator (LASSO), L1-regularized support vector machine (L1-SVM), and Random Forest. Features accumulated a total of 27,000 selection votes, and the 12 most stable features were retained.18-20 To identify the optimal classification algorithm, these candidate features were evaluated using K-nearest neighbors, support vector machine (SVM), and Random Forest classifiers. SVM demonstrated superior discriminatory performance and was selected for subsequent analyses.18,19,21

#### 3) Core Biomarker Identification and Model Evaluation

Core microbial biomarkers were defined as taxa identified by both machine learning feature selection (top 12 features) and univariate differential abundance analysis (FDR <0.05). Only overlapping taxa were retained for final model development. A reduced-feature SVM classifier was constructed using these core biomarkers and evaluated through nested cross-validation.^19^ The outer loop consisted of repeated stratified five-fold cross-validation, whereas hyperparameter optimization was performed using stratified three-fold cross-validation within the inner loop. Hyperparameters were selected by grid search, and the modeling pipeline incorporated feature standardization and balanced class weights. Model performance was evaluated using aggregated out-of-fold predictions. The area under the receiver operating characteristic curve (AUC) and corresponding bootstrap-derived 95% confidence intervals (CI) were calculated. Sensitivity and specificity were determined using the optimal threshold identified by the Youden index.

### Statistical Analysis

Continuous variables are presented as mean ± standard deviation or median (interquartile range), as appropriate. Categorical variables are reported as counts and percentages. Between-group comparisons were performed using the Mann–Whitney U test for continuous variables and Fisher’s exact test for categorical variables. A two-sided p value <0.05 was considered statistically significant.

## Results

### Baseline Characteristics

The study cohort comprised 33 STEMI patients, including 7 patients with primary VT/VF and 26 patients without primary VT/VF. Baseline demographic and clinical characteristics are summarized in Table 1. Age and body mass index (BMI) were comparable between groups. However, a significant sex imbalance was observed, with a higher proportion of female patients in the STEMI with VT/VF group than in the STEMI without VT/VF group (28.6% vs. 0%, p = 0.030). Cardiovascular comorbidities and in-hospital medical therapies did not differ significantly between groups. Although culprit vessel distribution was similar, STEMI patients with VT/VF experienced significantly longer ischemic times, reflected by a prolonged door-to-balloon time compared with patients without VT/VF (73.5 vs. 51.5 minutes, p = 0.003). Indicators of infarct severity, including left ventricular ejection fraction (LVEF), peak troponin I, and peak CK-MB levels, were comparable between groups. Serum potassium levels were numerically lower in the VT/VF group (3.4 vs. 3.7 mmol/L), although this difference did not reach statistical significance (p = 0.085).

**Table 1.** Baseline Demographic and Clinical Characteristics of STEMI Patients With and Without Primary VT/VF.

| Variables | STEMI with VT/VF<br>(n=7) | STEMI without VT/VF<br>(n=26) | *p* value |
| --- | --- | --- | --- |
| **Age** | 54.4 ± 13.8 | 56.7 ± 8.9 | 0.863 |
| **Male sex** | 5 (71.4%) | 26 (100%) | **0.030** |
| **Body mass index** | 24.9 ± 3.2 | 25.9 ± 3.1 | 0.415 |
| **Smoking** |  |  |  |
| Current smoker | 2 (28.6%) | 16 (61.5%) | 0.208 |
| Former smoker | 0 (0%) | 5 (19.2%) | 0.559 |
| **Comorbidities** |  |  |  |
| Hypertension | 5 (71.4%) | 9 (34.6%) | 0.106 |
| Diabetes mellitus | 1 (14.2%) | 9 (34.6%) | 0.397 |
| Ischemic stroke | 1 (14.2%) | 0 (0%) | 0.212 |
| Hyperlipidemia | 3 (42.9%) | 17 (65.4%) | 0.210 |
| CKD stage ≥ 3 | 2 (28.6%) | 3 (11.5%) | 0.281 |
| Prevalent atrial fibrillation | 1 (14.2%) | 0 (0%) | 0.212 |
| Previous heart failure | 0 (0%) | 1 (3.8%) | 1.000 |
| COPD | 0 (0%) | 0 (0%) | 1.000 |
| **Laboratory data** |  |  |  |
| Hemoglobin (g/dL) | 15.3 ± 1.1 | 13.7 ± 2.6 | 0.118 |
| White blood cell count (×10<sup>3</sup>/μL) | 11.8 ± 5.1 | 11.6 ± 4.0 | 1.000 |
| Platelet count (×10<sup>3</sup>/μL) | 263 ± 54 | 255 ± 66 | 0.826 |
| Total cholesterol (mg/dL) | 164.0 ± 54.5 | 179.9 ± 41.8 | 0.355 |
| Triglyceride (mg/dL) | 81.5 (75.0 – 90.2) | 114.0 (72.5 – 166.2) | 0.311 |
| LDL (mg/dL) | 113.0 ± 51.4 | 119.9 ± 39.1 | 0.582 |
| HDL (mg/dL) | 36.4 ± 7.4 | 41.5 ± 10.9 | 0.399 |
| Blood urea nitrogen (mg/dL) | 19.9 ± 16.3 | 16.6 ± 6.7 | 0.873 |
| Creatinine (mg/dL) | 1.25 ± 0.74 | 1.24 ± 0.55 | 0.725 |
| Sodium (mmol/L) | 142.6 ± 7.5 | 139.5 ± 1.6 | 0.531 |
| Potassium (mmol/L) | 3.4 ± 0.6 | 3.7 ± 0.5 | 0.085 |
| Alanine aminotransferase (U/L) | 83.0 (19.5 – 233.5) | 26.0 (22.0 – 35.0) | 0.261 |
| HbA1c (%) | 6.0 ± 0.2 | 6.7 ± 2.0 | 0.378 |
| Peak CK-MB (U/L) | 149.7 (90.8 – 261.6) | 150.6 (100.2 – 268.1) | 0.779 |
| Peak troponin I (ng/mL) | 15.9 (14.9 – 67.3) | 52.1 (31.7 – 120.9) | 0.323 |
| **LVEF (%)** | 53.2 ± 4.8 | 51.3 ± 9.1 | 0.838 |
| **Culprit Vessel<sup>a</sup>** |  |  |  |
| Left anterior descending | 4 (57.1%) | 10 (38.4%) | 0.422 |
| Left Circumflex | 1 (14.3%) | 3 (11.5%) | 1.000 |
| Right coronary artery | 2 (28.6%) | 14 (53.8%) | 0.398 |
| **Door-to-balloon time (min)** | 73.5 (58.5 – 92.2) | 51.5 (44.2 – 55.0) | **0.003** |
| **Post-PCI TIMI flow grade 3** | 7 (100%) | 26 (100%) | 1.000 |
| **Medications** |  |  |  |
| Aspirin + Clopidogrel | 1 (14.3%) | 0 (0%) | 0.212 |
| Aspirin + Ticagrelor/Prasugrel | 6 (85.7%) | 26 (100%) | 0.212 |
| Statin | 6 (85.7%) | 26 (100%) | 0.212 |
| **In hospital MACE** | 0 (0%) | 0 (0%) | 1.000 |

Abbreviations: CKD, chronic kidney disease; COPD, chronic obstructive pulmonary disease; LDL, low-density lipoprotein cholesterol; HDL, high-density lipoprotein cholesterol; HbA1c, glycated hemoglobin; CK-MB, creatine kinase–myocardial band; LVEF, left ventricular ejection fraction; PCI, percutaneous coronary intervention; TIMI, Thrombolysis In Myocardial Infarction; MACE, major adverse cardiovascular events.
Statistical significance was defined as $p < 0.05$ . Significant values are shown in bold italics.
<sup>a</sup>The cumulative percentage of culprit vessels exceeds 100% because one patient presented with simultaneous thrombosis of two coronary arteries (double culprit lesions).

### Microbial Diversity Analysis

Microbial diversity was evaluated at the species level across all study groups. Alpha-diversity analysis demonstrated significant differences in species richness as measured by the Observed (p = 0.032) and Chao1 (p = 0.019) indices (Figure 1). In contrast, Shannon and Simpson indices did not differ significantly among groups (all p > 0.05), indicating that changes in richness were not accompanied by substantial alterations in community evenness.

**Figure 1.**
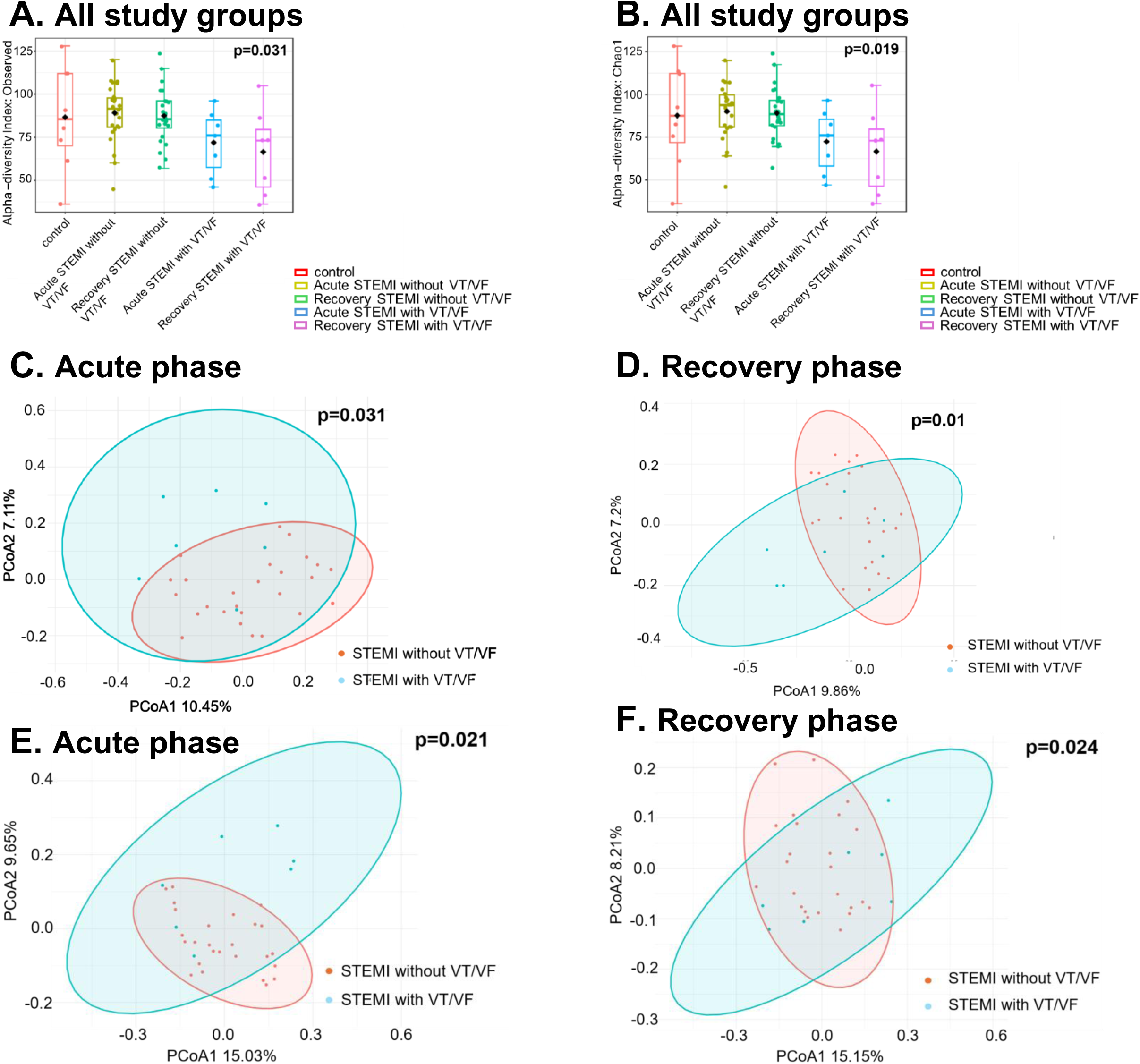
Alpha-diversity and beta-diversity measures across the five study groups. **(A)** Observed diversity in all study groups. **(B)** Chao1 index in all study groups. **(C)** Jaccard diversity between the Acute STEMI with and without VT/VF groups. **(D)** Jaccard diversity between the Recovery STEMI with and without VT/VF groups. **(E)** Unweighted UniFrac diversity between the Acute STEMI with and without VT/VF groups. **(F)** Unweighted UniFrac diversity between the Recovery STEMI with and without VT/VF groups.

Beta-diversity analysis revealed significant compositional differences between STEMI patients with and without primary VT/VF in both the acute and recovery phases when assessed using presence/absence-based metrics. Significant separation was observed using unweighted UniFrac distances (acute phase: p = 0.021; recovery phase: p = 0.024) and Jaccard distances (acute phase: p = 0.031; recovery phase: p = 0.002) (Figure 1). In contrast, abundance-weighted metrics, including weighted UniFrac and Bray–Curtis dissimilarity, did not demonstrate significant clustering between groups.

### Taxonomic Differential Abundance Analysis

LEfSe analysis identified marked differences in microbial composition between STEMI patients with and without primary VT/VF. During the acute phase, the VT/VF group exhibited enrichment of 25 species, 7 genera, 3 families, and 1 order (LDA score >2.0; p < 0.05) (Figure 2). During the recovery phase, 15 species and 5 genera remained significantly enriched in the primary VT/VF group.

**Figure 2.**
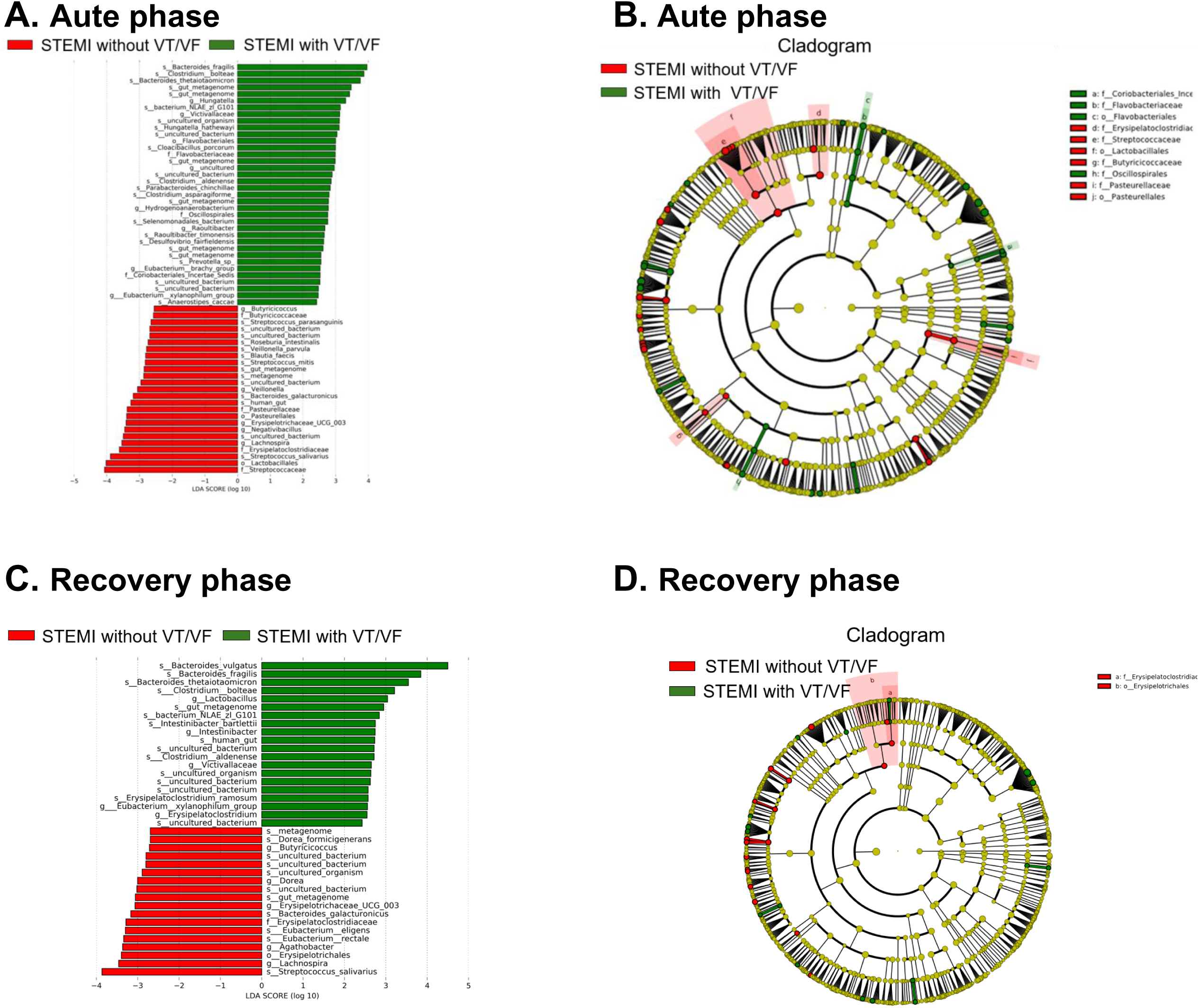
Differentially abundant taxa identified by linear discriminant analysis (LDA) effect size (LEfSe) and taxonomic cladogram. LEfSe was performed on stool microbiome profiles to identify taxa enriched among the STEMI without VT/VF and STEMI with VT/VF groups. **(A)** Histogram of LDA scores showing differentially abundant bacterial taxa comparing acute STEMI with and without VT/VF group (LDA score > 2.0). **(B)** Taxonomic cladogram summarizing the phylogenetic distribution of significantly enriched taxa in the acute phase. **(C)** Histogram of LDA scores comparing recovery STEMI with and without VT/VF group. **(D)** Corresponding cladogram highlighting differentially abundant taxa in the recovery phase. In all panels, green and red indicate taxa enriched in STEMI with and without VT/VF groups, respectively.

Among the most prominent taxa enriched during the acute phase were *Bacteroides fragilis*, *Enterocloster bolteae* (formerly *Clostridium bolteae*), and *Bacteroides thetaiotaomicron*. Notably, *Bacteroides fragilis* and *Bacteroides thetaiotaomicron* remained enriched during the recovery phase, together with *Phocaeicola vulgatus* (formerly *Bacteroides vulgatus*), suggesting persistence of these microbial alterations beyond the acute STEMI period.

### Machine Learning–Based Feature Selection

For the primary classification task comparing acute STEMI patients with and without primary VT/VF, the dataset included 33 subjects, 741 microbial features, and 59 clinical variables. A bootstrap-based feature selection framework integrating LASSO, L1-SVM, and Random Forest algorithms identified 12 highly stable predictors.

The most consistently selected features included serum creatinine, *Alistipes shahii*, three *Lachnoclostridium*-related taxa (*Clostridium aldenense*, *Enterocloster bolteae*, and *bacterium NLAE-zl-G101*), two *Roseburia* taxa, *Ruminococcus lactaris* (*R. torques* group), and a *Klebsiella pneumoniae*–related taxon. Although serum creatinine was frequently selected by the machine learning framework, it did not differ significantly between groups in univariate analysis (p = 0.974), suggesting that its predictive value was driven primarily by multivariable interactions.

In the recovery-phase analysis, feature-selection stability was substantially lower, with only a limited number of taxa—including Intestinibacter bartlettii, Subdoligranulum spp., Lachnoclostridium taxa, and Bacteroides fragilis—consistently retained across bootstrap iterations (data not presented). Given the reduced stability of recovery-phase predictors, no further classification modeling was pursued.

### Core Biomarker Identification and Model Performance

To identify robust microbial biomarkers associated with primary VT/VF, the top 12 machine learning–derived features were intersected with taxa demonstrating significant differential abundance by edgeR (FDR <0.05). Five taxa fulfilled both criteria (Figure 3):

1. *Clostridium aldenense*
2. *Enterocloster bolteae*
3. *Lachnoclostridium bacterium NLAE-zl-G101*
4. *Alistipes shahii*
5. *Roseburia* sp.

**Figure 3.**
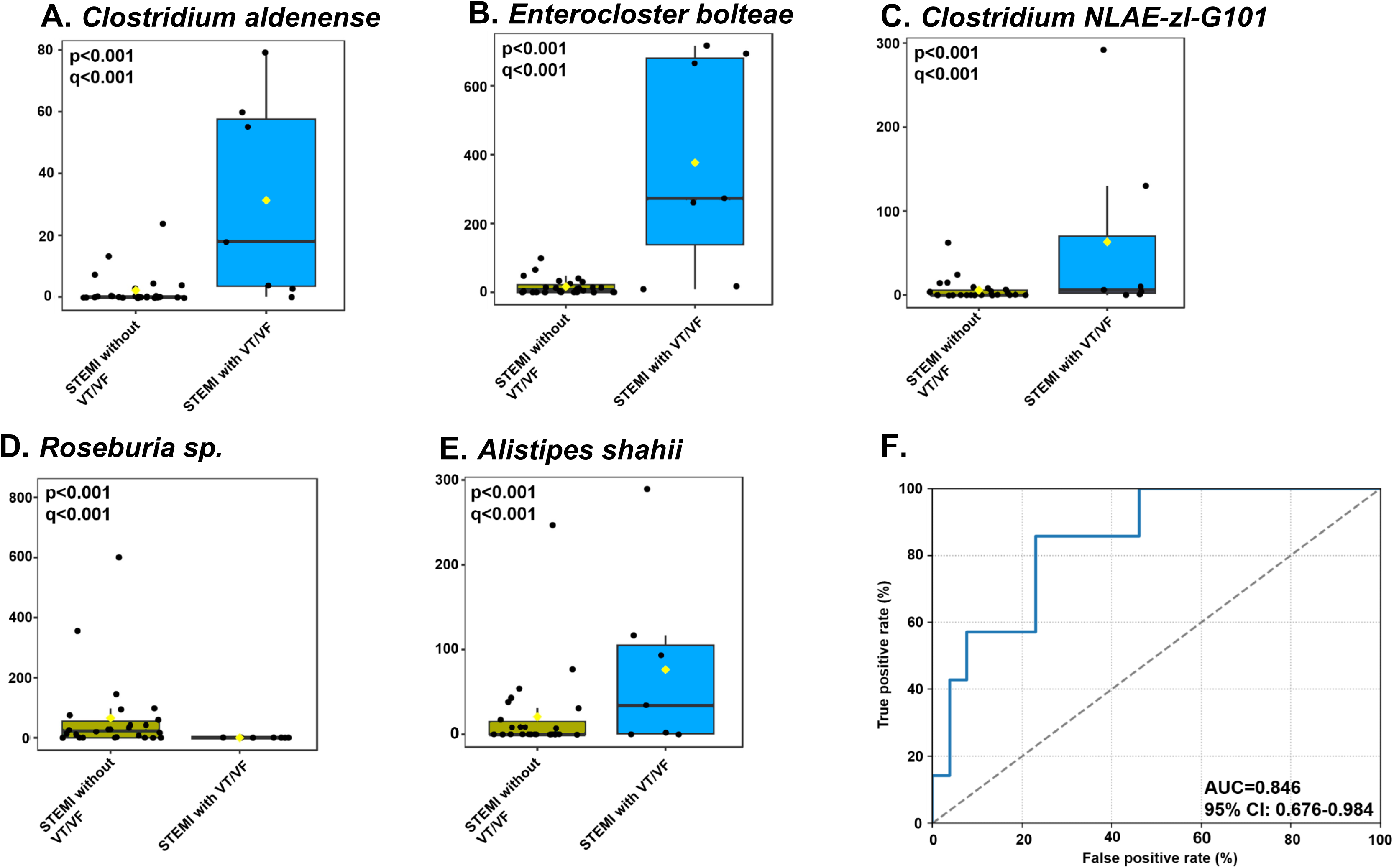
Differential abundance of specific microbial taxa and their diagnostic performance. **(A)**-**(E)** comparison of filtered counts for five specific taxa. **(A)** *Clostridium aldenense*, **(B)** *Enterocloster bolteae*, **(C)** *Clostridium NLAE-zl-G101,* **(D)** *Roseburia sp*, and **(E)** *Alistipes shahii*. (**F)** Receiver Operating Characteristic (ROC) curve generated using a Support Vector Machine (SVM) model based on the five aforementioned taxa.

These five taxa were designated as core microbial biomarkers associated with primary VT/VF during the acute phase of STEMI. A reduced-feature SVM model constructed using only these biomarkers achieved an area under the receiver operating characteristic curve (AUC) of 0.846 (95% CI: 0.676–0.984), with a sensitivity of 85.7% and a specificity of 76.9% for distinguishing STEMI patients with primary VT/VF from those without VT/VF (Figure 3).

### Functional Metabolic and Enzymatic Profiling

Functional analyses were restricted to acute-phase samples because recovery-phase taxa did not demonstrate sufficient stability for robust machine learning–based discrimination. PICRUSt2 identified 13 metabolic pathways that differed significantly between STEMI patients with and without primary VT/VF **(Supplementary Figure 1)**. Compared with the non-VT/VF group, the VT/VF group demonstrated significant enrichment of pathways involved in mucin-derived carbohydrate metabolism, including the superpathway of fucose and rhamnose degradation (p = 0.002) and fucose degradation (p = 0.015). In addition, guanosine nucleotide degradation III was significantly enriched (p = 0.020). Conversely, the mono-trans, poly-cis decaprenyl phosphate biosynthesis pathway was significantly depleted in the primary VT/VF group (p = 0.008) (Figure 4).

**Figure 4.**
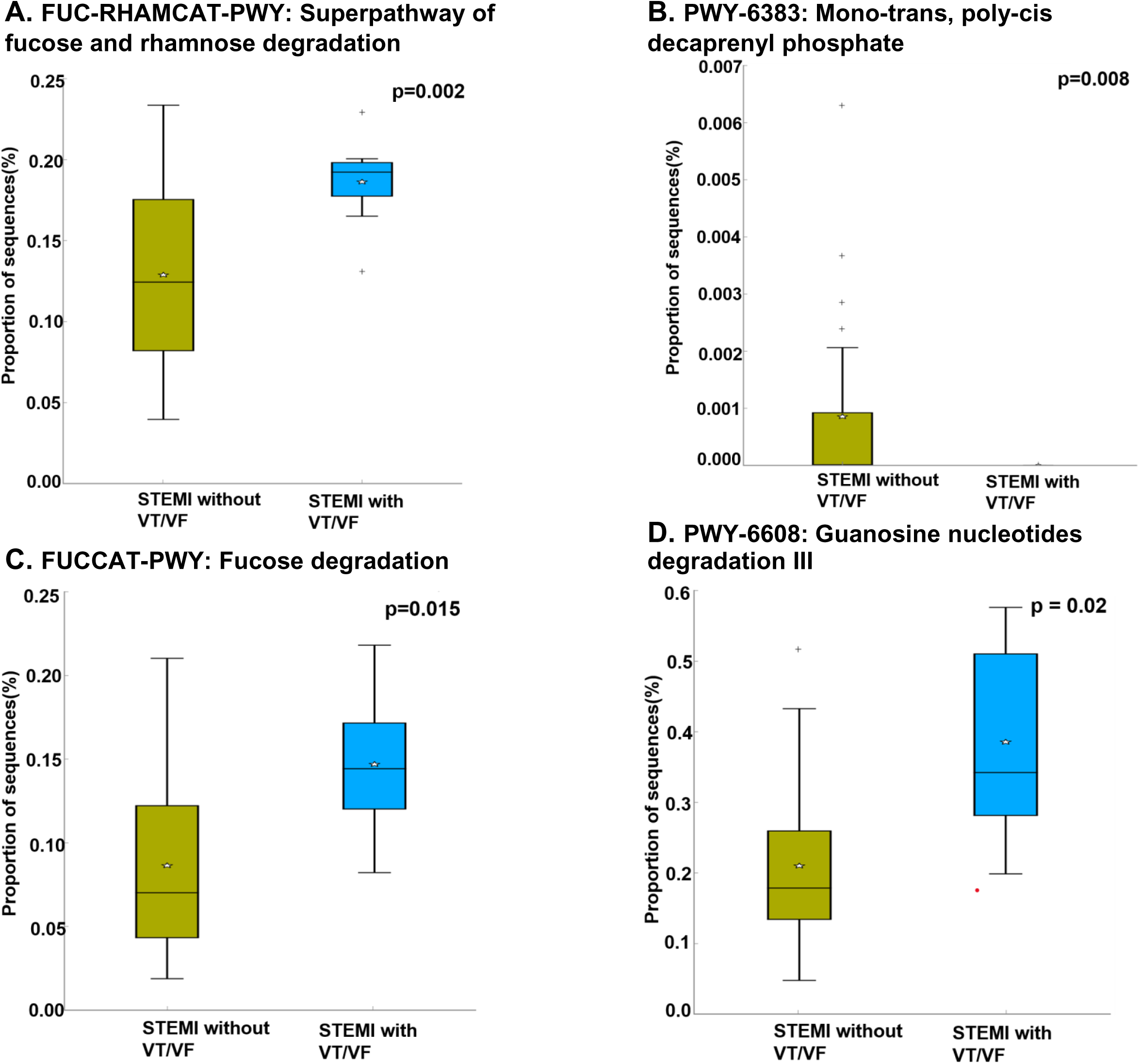
Comparison of the relative abundance of predicted metabolic pathways between acute STEMI patients with and without VT/VF. Boxplots display the proportion of sequences for pathways exhibiting significant differences between the two groups. The vertical axis indicates the percentage of sequences, and statistical significance is denoted by P-values.

At the enzymatic level, 115 enzymes differed significantly between groups **(Supplementary Figure 2)**. The primary VT/VF group exhibited reduced abundances of (2Z,6E)-farnesyl diphosphate synthase (EC 2.5.1.68, p = 0.008) and undecaprenyl-diphosphate phosphatase (EC 3.6.1.27, p = 0.026), together with increased abundances of L-fucose mutarotase (EC 5.1.3.29, p = 0.033) and xanthine dehydrogenase (EC 1.17.1.4, p = 0.047) (Figure 5). Collectively, these findings suggest enhanced microbial fucose utilization and purine catabolism in STEMI patients who develop primary VT/VF. Notably, these functional alterations were concordant with the enrichment of specific VT/VF-associated taxa, including *Bacteroides fragilis*, and *Bacteroides thetaiotaomicron*, supporting the presence of a coordinated gut microbiome–metabolic signature associated with arrhythmogenic susceptibility in STEMI.^22,23^

**Figure 5.**
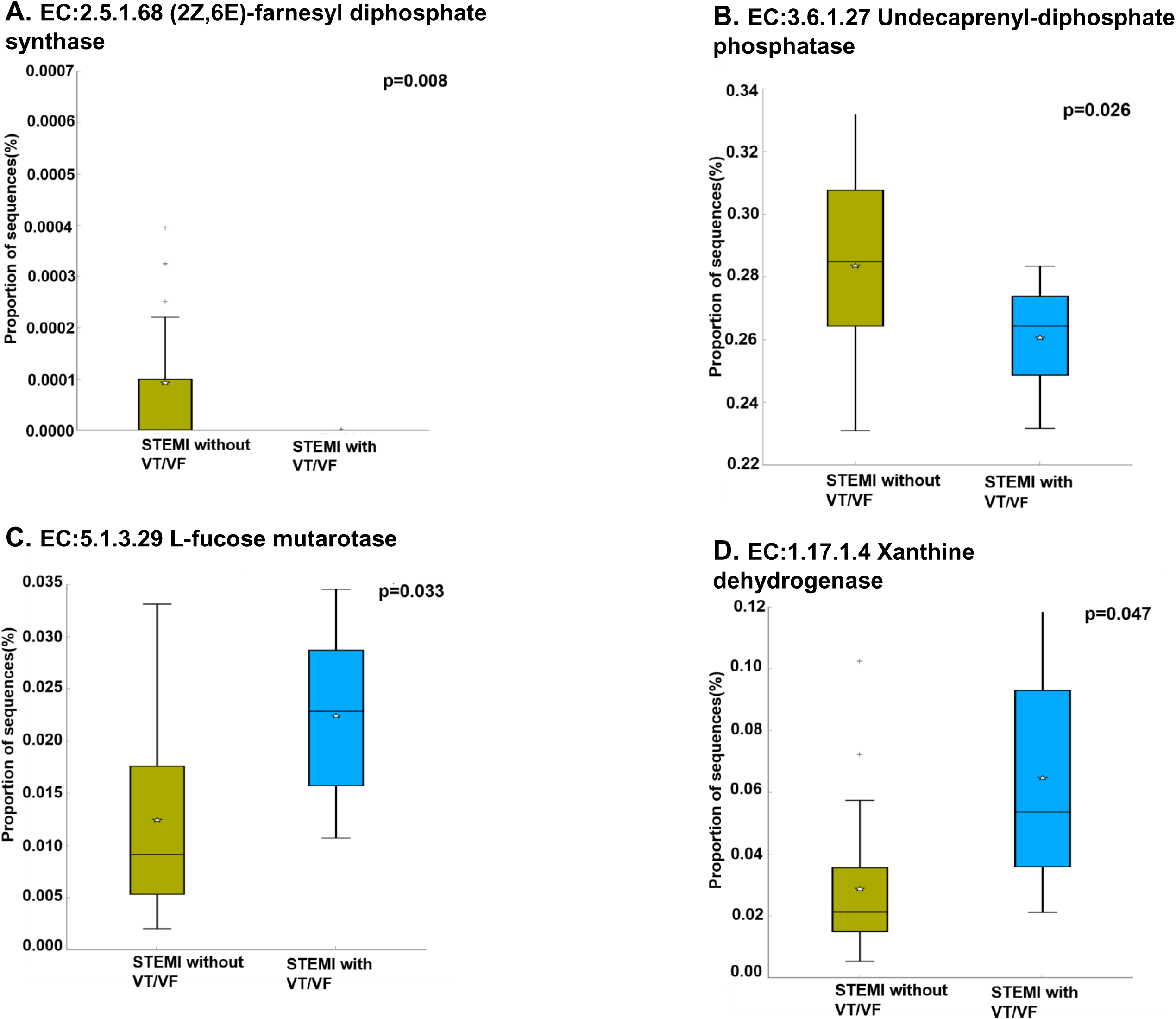
Comparison of the relative abundance of predicted microbial enzymes between acute STEMI patients with and without VT/VF. Specific EC numbers and enzyme names are indicated above each plot. The vertical axis indicates the percentage of sequences, and statistical significance is denoted by P-values.

## Discussion

To our knowledge, this is among the first studies to characterize gut microbiome alterations in STEMI patients presenting with primary VT/VF using an integrated machine learning and microbiome profiling approach. The principal finding was the identification of a distinct microbial signature comprising three *Lachnoclostridium*-related taxa (*Clostridium aldenense*, *Enterocloster bolteae*, and *bacterium NLAE-zl-G101*), *Alistipes shahii*, and *Roseburia* sp. These taxa consistently emerged through both machine learning and differential abundance analyses, and a reduced-feature support vector machine model based on these biomarkers achieved an internally validated AUC of 0.846. In addition, functional profiling revealed coordinated alterations in microbial carbohydrate and purine metabolism, supporting the presence of a gut microbiome–metabolic signature associated with VT/VF in STEMI.

### Alterations in Gut Microbial Diversity and Community Structure

Alpha-diversity analysis demonstrated significant differences in species richness across study groups, as reflected by the Observed and Chao1 indices. Although formal post-hoc testing was not performed, the numerically lower richness observed in STEMI patients with primary VT/VF is broadly consistent with previous reports describing reduced microbial diversity in patients with acute myocardial infarction compared with healthy controls.^24^ Given the exploratory nature of these analyses and differences in study populations, these findings should be interpreted cautiously.

Beta-diversity analyses revealed significant separation between STEMI patients with and without primary VT/VF when assessed using unweighted UniFrac and Jaccard distances, whereas abundance-weighted metrics did not demonstrate significant clustering. This pattern suggests that the observed differences were driven primarily by low-abundance taxa rather than dominant community members.

### Machine Learning–Derived Microbial Predictors of Primary VT/VF

A central finding of this study was the identification of a microbial signature enriched in STEMI patients who developed primary VT/VF. Three members of the *Lachnoclostridium* lineage consistently emerged as core biomarkers across analytical approaches. The most striking finding of this study was the consistent enrichment of three *Lachnoclostridium*-related taxa (*Clostridium aldenense*, *Enterocloster bolteae*, and *bacterium NLAE-zl-G101*) in STEMI patients with primary VT/VF. Despite differing levels of biological characterization, all three taxa belong to a microbial lineage increasingly associated with cardiometabolic disease and inflammatory dysbiosis. Their concurrent enrichment, together with predicted activation of fucose degradation and purine catabolic pathways, suggests that a coordinated Lachnoclostridium-associated metabolic niche may contribute to the gut microbiome– metabolic signature linked to arrhythmogenic susceptibility during acute STEMI.

*Alistipes shahii* was also identified as a core biomarker by machine learning–based framework despite not reaching the LEfSe significance threshold. Previous studies have linked *Alistipes* species to alterations in host lipid metabolism and systemic fatty acid profiles.^25,26^ Elevated long-chain fatty acid concentrations have been shown experimentally to impair cardiac sodium and potassium currents and disrupt gap-junction coupling, thereby increasing conduction heterogeneity and susceptibility to reentrant arrhythmias. ^27–29^ Although direct metabolomic measurements were not performed in the present study, these observations suggest a plausible mechanistic link between *Alistipes* enrichment and arrhythmogenic risk.

In contrast, *Roseburia* species are well-established butyrate producers that contribute to intestinal barrier integrity and immune homeostasis ^30,31^ Depending on the directionality observed in the differential abundance analyses, alterations in *Roseburia* abundance may reflect disruption of protective microbial functions during the acute phase of STEMI complicated by primary VT/VF. Because 16S rRNA sequencing provides limited functional resolution, further metagenomic and metabolomic studies are required to clarify the biological role of these taxa in arrhythmogenesis.

### Alterations in Microbial Metabolic Pathways and Enzymatic Profiles

Functional prediction analyses revealed enrichment of pathways involved in fucose and rhamnose degradation among STEMI patients with primary VT/VF. These pathways are closely linked to microbial utilization of host-derived glycans and have previously been associated with intestinal responses to inflammation, hypoxia, and epithelial stress.^32,33^ Complementing the machine learning–derived microbial signature, LEfSe analysis demonstrated that *Bacteroides fragilis* and *Bacteroides thetaiotaomicron* remained enriched during the recovery phase, together with *Phocaeicola vulgatus*, suggesting that certain microbiome alterations persist beyond the acute STEMI event. This temporal persistence indicates that the observed dysbiosis may reflect a sustained microbial remodeling process rather than a transient response to acute ischemic injury. Notably, *Bacteroides thetaiotaomicron* is a well-characterized mucin-degrading bacterium with the capacity to utilize host-derived fucosylated glycans.^22^ Its enrichment is concordant with the predicted activation of fucose degradation pathways identified by PICRUSt2, providing biological coherence between the taxonomic and functional analyses. Together, these findings support the presence of a coordinated gut microbiome–metabolic signature associated with primary VT/VF in STEMI patients. Furthermore, increased abundance of L-fucose mutarotase, a key enzyme involved in fucose metabolism, further supported activation of this metabolic pathway.

In addition to carbohydrate metabolism, we observed significant alterations in microbial purine metabolism. The primary VT/VF group demonstrated enrichment of guanosine nucleotide degradation pathways together with increased predicted abundance of xanthine dehydrogenase. Although the specific microbial contributors to these functional changes cannot be determined from 16S rRNA sequencing alone, the concurrent enrichment of *Lachnoclostridium*-related taxa and persistent overrepresentation of *Bacteroides fragilis* and *Bacteroides thetaiotaomicron* suggest broader microbial community remodeling associated with altered metabolic capacity. Xanthine dehydrogenase and its oxidase form catalyzed the conversion of purines to uric acid while generating reactive oxygen species. Previous studies have implicated xanthine oxidase–derived oxidative stress in ischemia–reperfusion injury and cardiac arrhythmogenesis.^34,35^ Although these findings are based on computational functional prediction rather than direct enzyme activity measurements, the concordance between taxonomic alterations and predicted enrichment of purine catabolism supports the presence of a coordinated gut microbiome–metabolic signature associated with ventricular electrical instability in STEMI patients who develop primary VT/VF.

Together, the enrichment of *Lachnoclostridium*-related taxa, persistence of *Bacteroides* species with established glycan-utilization capabilities, activation of fucose degradation pathways, and increased predicted purine catabolism collectively support the existence of a coordinated gut microbiome–metabolic signature associated with arrhythmogenic susceptibility in STEMI patients presenting with primary VT/VF. Future multi-omics studies integrating metagenomics, metabolomics, and mechanistic validation will be necessary to determine whether these microbial alterations are causal contributors to primary VT/VF or biomarkers of disease severity.

## Limitations

This study has several limitations. First, the sample size was relatively small, particularly in the STEMI with primary VT/VF group, which may limit statistical power and increase the risk of model overfitting. Although the machine learning framework incorporated bootstrap-based feature selection and nested cross-validation, the resulting classification model should be considered exploratory and requires validation in larger independent cohorts before clinical application.

Second, a significant sex imbalance was present between groups, with all patients in the STEMI without VT/VF group being male. Although STEMI occurs more frequently in men, this imbalance may introduce sex-related confounding and limit the generalizability of the observed microbial signatures. Future studies with more balanced sex distributions are needed to evaluate potential sex-specific microbiome effects.

Third, the observational design precludes causal inference. Because stool samples were collected after the acute ischemic event, it remains uncertain whether the identified microbial alterations represent pre-existing pro-arrhythmic factors, adaptive responses to myocardial injury, or secondary consequences of the systemic inflammatory and hemodynamic stress associated with STEMI and primary VT/VF.

Fourth, dietary habits, lifestyle factors, and medication exposures that may influence gut microbial composition were not systematically recorded. Although recruitment from a geographically localized population may have reduced some environmental heterogeneity, residual confounding cannot be excluded.

Finally, functional pathway analyses were based on computational inference from 16S rRNA sequencing data rather than direct metagenomic, transcriptomic, proteomic, or metabolomic measurements. Consequently, the predicted alterations in fucose metabolism and purine degradation should be regarded as hypothesis-generating and require validation through multi-omics approaches and mechanistic studies.

## Conclusions

This study identified a distinct gut microbiome–metabolic signature associated with primary VT/VF in STEMI, characterized by enrichment of *Lachnoclostridium*-related taxa during the acute phase and persistent overrepresentation of *Bacteroides fragilis* and *Bacteroides thetaiotaomicron* across both the acute and recovery phases. These taxonomic alterations were accompanied by enhanced microbial fucose degradation and purine catabolic pathways, supporting a coordinated gut microbiome–metabolic remodeling associated with arrhythmogenic susceptibility. Collectively, these findings provide a rationale for exploring microbiome-derived biomarkers and metabolic pathways as future tools for precision risk stratification and mechanistic investigation of primary VT/VF in STEMI.

## Non-standard Abbreviations and Acronyms

ASV: amplicon sequence variant
CCS: circular consensus sequencing
CLR: centered log-ratio
FDR: false discovery rate
LDA: linear discriminant analysis
LEfSe: linear discriminant analysis effect size
ML: machine learning
PICRUSt2: Phylogenetic Investigation of Communities by Reconstruction of Unobserved States 2
SVM: support vector machine
VT/VF: ventricular tachycardia/ventricular fibrillation

## Acknowledgments

The authors thank the study participants and the clinical and laboratory staff who supported sample collection and processing.

## Previous Presentation

This study was presented in part at the Heart Rhythm Society Annual Meeting 2026, Chicago, Illinois, USA.

